# FIS103, a Novel SULT1A1-dependent Prodrug, Demonstrates Potent Antitumor Activity in Renal Cell Carcinoma

**DOI:** 10.1101/2024.03.21.24304257

**Authors:** Ross A. Hamilton, Uksha Saini, Mai Tran, Christopher J. Foley, Pooja Enagala, Leniher C. Chibas, Disha Chatterjee, Stephanie P. Vega, Dev Chatterjee, Atul Varadhachary

## Abstract

Intra-tumoral heterogeneity has been shaping the field of precision medicine for cancer patients ever since its emergence. Prodrugs, which require activation by tumor associated enzymes (TAEs), are a rapidly emerging approach for targeted therapeutics. SULT1A1, a sulfotransferase enzyme and TAE, is over-expressed in about 5-15% of cancer patients including breast, prostate and renal cell carcinoma (RCC); however, it is either not expressed or expressed at low level in most normal tissue. Bioinformatic RNA analyses revealed that SULT1A1 over-expression in tumors is correlated with worse patient prognosis. We have identified a new compound, FIS103, which is a small molecule anti-cancer prodrug that is activated by SULT1A1 once internalized. This class of compounds, N-benzyl indole carbinols (N-BICs), cause rapid cell death by inducing widespread non-specific covalent alkylation of proteins in the cancer cell. We report that FIS103 displays potent antitumor activity in SULT1A1 over-expressing RCC cell lines (A498 and Caki-1). Contrarily, low SULT1A1 expressing RCC cells (786-O and ACHN) did not show any antitumor effects, which suggests low FIS103 toxicity in the absence of SULT1A1. *In silico* modeling validated the predicted SULT1A1-FIS103 interaction. Furthermore, FIS103 demonstrates potent SULT1A1-dependent antitumor activity in NU/J mouse xenografts injected with A498 cells. Remarkably, the flank tumors in mice regressed to non-measurable 14 days post-FIS103 treatment and did not regrow through the study conclusion. Additionally, SD rats treated with FIS103 once daily for 14 days demonstrated a promising liver toxicity profile with serum liver enzymes falling within the normal range and histopathology analysis indicated no difference between FIS103 or vehicle treated rats. We hereby demonstrate that FIS103 may have the potential to improve survival as well as quality of life of RCC patients and its application could be extended to other SULT1A1 expressing cancers.

## Introduction

Improved characterization of the etiology and complex biology of cancer cells has led to the development of precision medicines, reliant on molecules prevalent in cancer cells to exert therapeutic effects while leaving healthy tissue unaffected. Recently, developments have focused on designing intracellular targeted chemotherapeutics with site-specific delivery of cytotoxic anticancer agents. One strategy is the delivery of prodrugs activated by tumor-associated enzymes (TAEs), a molecular biomarker. TAEs are present at elevated levels in tumor tissue; examples include extracellular enzymes in necrotic areas within tumors (e.g., beta-glucuronidase[1]) and tumor-associated proteases in invasive and metastatic tumors (e.g., plasmin[2]). Currently, there are several prodrugs utilizing this strategy being developed for targeted cancer therapies. The most common targets are lysosomal proteases (e.g, cathepsin[3]) and proteases in the extracellular matrix (e.g, matrix metalloprotease[4, 5]). Designing selective prodrugs activated only by TAEs has significant potential for cancer therapeutics due to their specific targeting of tumor cells with minimal cytotoxicity to healthy tissues.

SULT1A1 is an enzyme belonging to the family of sulfotransferase enzymes that utilize 3’-phospho-5’-adenylyl sulfate (PAPS) to catalyze sulfate conjugation of numerous acceptor molecules bearing a hydroxyl or an amine group[6, 7]. This modification predominantly increases its solubility and decreases its biological activity. It metabolizes several phenolic substrates including simple phenolic compounds[8], drugs (e.g., acetaminophen[9], minoxidil[10]); estrogens (e.g., β-estradiol[11]), and synthetic estrogenic compounds (e.g., trans-4-hydroxytamoxifen[12, 13]). The enzyme is primarily expressed in the gastrointestinal tract, specifically in certain stomach, liver, duodenum, small intestine, colon, rectum and appendix cell types[14, 15] (Human Protein Atlas).

SULT1A1 is also known to bioactivate procarcinogens including N-hydroxy metabolites of 2-amino-3-methylimidazo[4,5-f]quinoline[16] and other procarcinogens[17]. This metabolic activation of carcinogenic N-hydroxyarylamines produces highly reactive intermediates capable of forming DNA adducts, potentially resulting in mutagenesis[18]. SULT1A1 was discovered to be a marker for certain carcinogen-induced malignancies[19]. Abnormal SULT1A1 expression has been reported in some cancers even when it is absent in adjacent normal tissue (e.g., breast cancer), and in other cancers with increased expression levels (e.g., hepatocellular carcinoma)[13, 14, 20]. 8-15% of these malignancies show high SULT1A1 expression (Table 1). Additionally, SULT1A1 has been implicated as an oncogene and a negative prognostic marker for many cancers, including non-small cell lung cancer (NSCLC), pediatric neuroblastoma, and renal cell carcinoma (RCC)[21-23]. These cancer subtypes are fairly aggressive and prevalent; NSCLC has a 5-year survival rate of 26% and around 236,740 adults were estimated to be diagnosed with NSCLC in 2022 (American Cancer Society).

**Table 1.** Percent of patients with high (>1.3 Z-scores) SULT1A1 mRNA expression (TCGA datasets)

| Cancer Type | % Patients w/<br>High SULT1A1 | New Diagnoses<br>/yr | Potential New Patients<br>for Treatment / yr |
| --- | --- | --- | --- |
| Breast | 9% | 225,180 | 22,966 |
| Lung (NSCLC) | 8% | 189,125 | 15,130 |
| Brain (GBM) | 9% | 12,376 | 1,113 |
| Prostate | 15% | 161,360 | 24,204 |
| Kidney (CCC) | 9% | 58,870 | 5,298 |
| Pancreatic | 10% | 53,670 | 5,367 |
| Liver (HCC) | 8% | 30,532 | 2,442 |
| Total |  |  | 76,520 |

One class of antitumor compounds, N-benzyl indole carbinols (N-BICs), has been shown to function as prodrugs activated by SULT1A1. N-BICs are synthetic derivatives of a natural product, indole-3-carbinol (I3C), which is found in cruciferous vegetables and thought to have anticancer properties[24-27]. N-BICs differ from their I3C parent by the benzyl group off of their indole nitrogen, which gives them greater chemical stability in acidic conditions[28, 29]. N-BICs exert their cytotoxic effect by non-specific covalent alkylation of proteins, resulting in rapid cell death and tumor suppression[30]. It is thought that N-BICs require the expression of SULT1A1 for antitumor activity[31, 32]. The published *in vitro* and *in vivo* toxicity of N-BIC compounds in cancer cells specifically expressing SULT1A1 provides scientific proof-of-concept supporting the development of these compounds. N-BICs have also demonstrated potent anti-tumor efficacy in several solid and hematologic tumors[33-36].

*In silico* prediction software generated new chemical analog compound classes based on the N-BIC core molecule to capitalize on TAE expression to develop improved precision therapies. The N-BIC analogs designed at this stage were expected to retain or improve anti-tumor activity, as well as increase solubility and decrease sensitivity to acidic conditions. 20 total analogs were created with the best predicted performances across the metrices; of those, FIS103 showed differential cell killing consistent with SULT1A1 expression. These were derived from NSC-743380 (parental compound), an N-BIC analog designed for anti-leukemia activity in SULT1A1-expressing acute myeloid leukemia cells[32]. Our clinical candidate exhibited excellent cytotoxic efficacy in RCC models expressing SULT1A1 *in vitro* and *in vivo*, and has encouraging pharmaceutical properties like aqueous solubility and stability, and a promising liver toxicity profile. Since SULT1A1 has not yet been explored as a target for prodrugs, FIS103 is a novel therapy with great potential upside for combatting RCC and multiple other aggressive cancer subtypes where SULT1A1 expression is prevalent.

## Materials and Methods

### Bioinformatics Analyses of SULT1A1 Expression

Using SULT1A1 RNA-seq expression data, we undertook a pan-cancer classification of The Cancer Genome Atlas (TCGA) tumor samples. Patients with >1.3 Z-score were considered to have high mRNA expression levels of SULT1A1. All TCGA data was extracted via cBioPortal. New diagnoses / year statistics from Cancer Facts and Figures[37]. Kaplan-Meier overall survival curves were generated from TCGA data using SurvivalGenie, a platform for survival analysis across pediatric and adult cancers[38].

### *In Silico* Prediction of N-Bic Analogs for Enhanced Anti-tumor Activity

In collaboration with American Biochemicals, N-BIC analogs based on the core molecule were generated using *in silico* prediction software. To further screen compounds, AI software assessed proposed pharmaceutical properties of absorption, distribution, metabolism, excretion, and toxicity (ADMET). The ADMET Predictor software analyzes each analog’s chemical, pharmacokinetic, and toxicological properties, including LogP, pH/pKa, and metabolism (Figure 2B).

**Figure 1.**
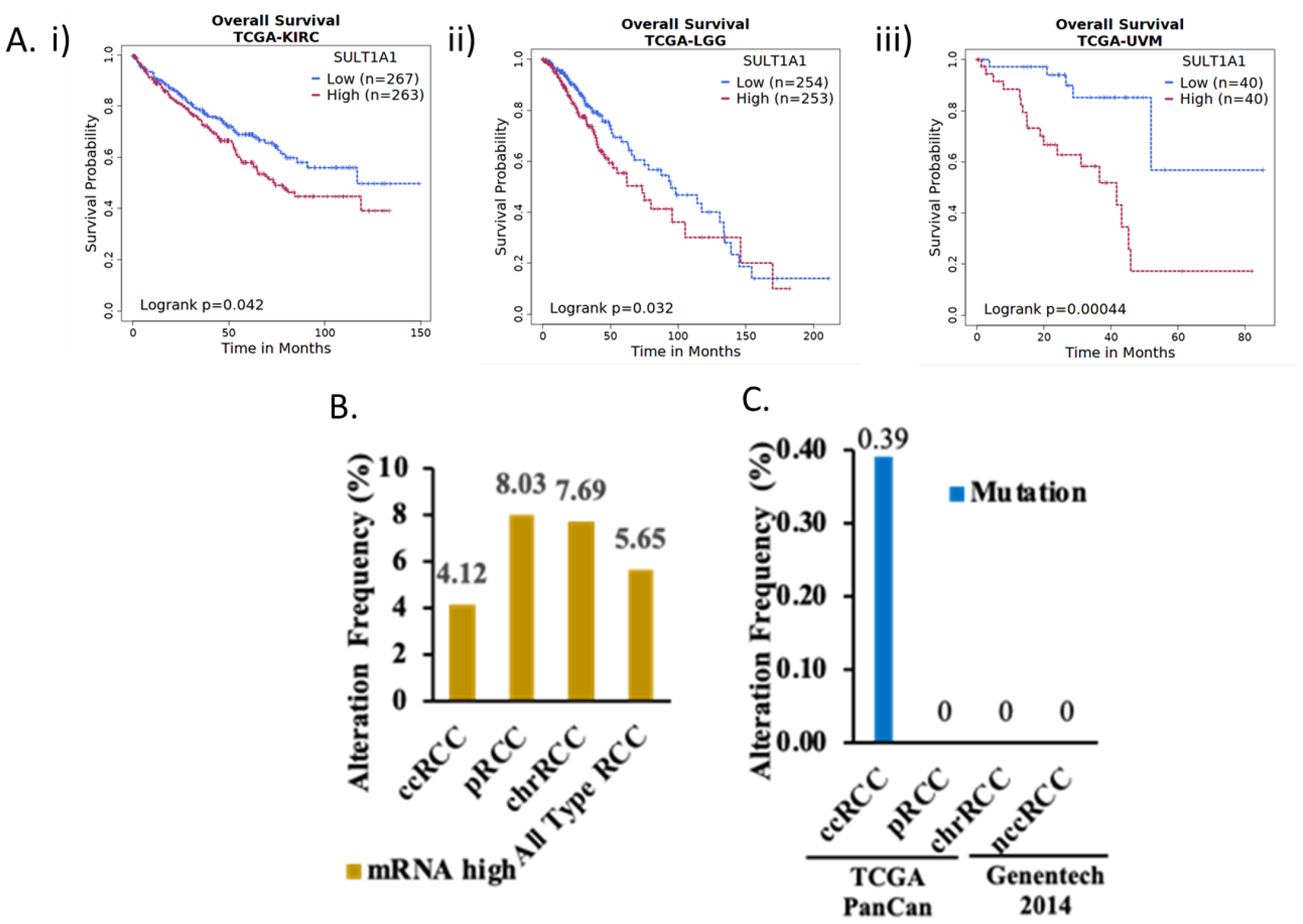
SULT1A1 expression correlates with poorer patient outcomes in specific cancers. A) Kaplan-Meier Plots indicating that high expression of SULT1A1 mRNA correlates with poor prognosis for patients with i) kidney renal clear cell carcinoma, ii) brain lower grade glioma, and iii) uveal melanoma. Percentage of patients with B) high SULT1A1 mRNA expression in RCC subtypes (RNASeq from the TCGA PanCancer study), and C) with SULT1A1 mutation (whole genome sequencing from the TCGA PanCancer study and Genentech 2014 study). All TCGA data was extracted via cBioPortal. The number of cases per RCC subtypes is as followed: clear cell RCC (ccRCC) n=512, papillary RCC (pRCC) n=283, chromophobe RCC (chrRCC) n=65, non-clear cell RCC (nccRCC) n=146.

**Figure 2.**
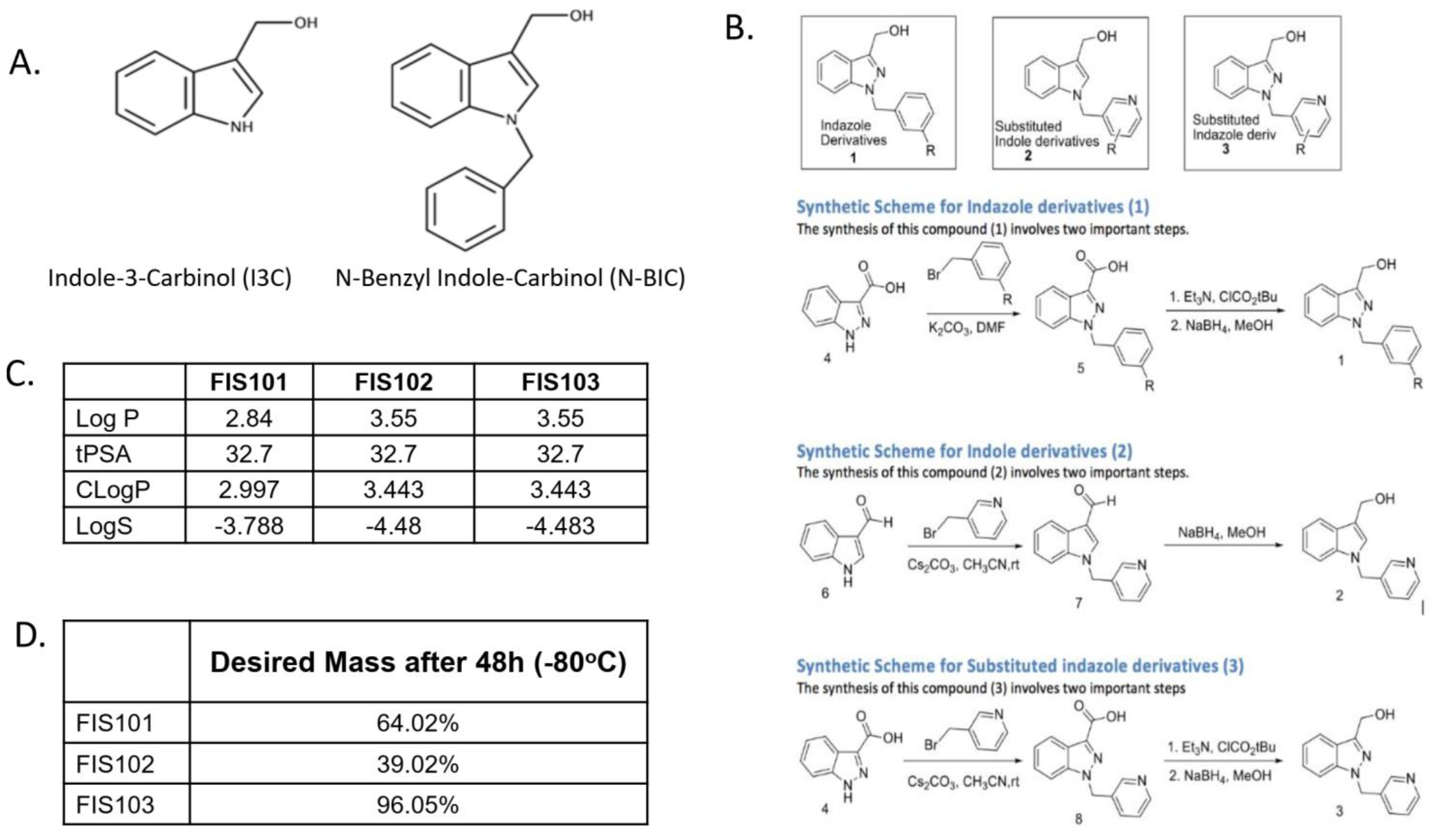
Prediction and synthesis of N-BIC analogs. A) Indole-3-Carbinol (I3C) and N-BIC core compound structures. B) Preliminary schema for N-BIC analogs proposed by American Biochemical. C) Performance of the 3 lead compounds from *in silico* analysis. D) Stability of three generated analogs of N-BIC core compound. Percentage represents liquid chromatography mass spectrometry trace.

### Synthesis of N-BIC Analogs

Further *in silico* analysis and molecular design was conducted in collaboration with JRF International. The computational analysis was based on *in silico* analyses of each analog’s aqueous solubility, predicted volume of distribution, and native pH level for advancement to synthesis and functional testing. To improve compound solubility to desired levels, charged or polar chemical groups such as methoxy, amino groups (of moderate acid sensitivity), benzoic acid, or sulfonic acid were added. In total, 20 N-BIC analogs were generated; of these, FIS101, FIS102, and FIS103 were selected for synthesis.

### *In Silico* Prediction of FIS103 Binding to SULT1A1

Studies were conducted by the Texas Medical Center’s Accelerator for Cancer Therapeutics (ACT). Protein Data Bank (PDB) entries of SULT1A1 crystal structures (PDB IDs: 2D06, 1LS6, 3QVU, 3QVV, 3U3K, 3U3M, and 3U3O) were superimposed, and their conformational variability at known SULT1A1 substrate binding sites was analyzed. Comparison between PDB IDs 2D06 and 1LS6 showed residue displacement; hence, both files were chosen for structure-based virtual screening. To dock FIS103, The software Gromacs v2020 was used because it permits for flexible ligand conditions, allowing compound shifts to accommodate changes in the binding pocket[39]. To generate a binding site model for FIS103 and compare the binding mode to that of p-nitrophenol or estradiol (known SULT1A1 substrates), FIS103 was docked at the defined grid using the automated “standard precision” (SP) mode of Glide. To further confirm this, the top scoring docking model for 2D06 and 1LS6 was submitted to a short 100 ns MD simulations.

### Cell Lines and Media

Cell lines (sourced from ATCC) were incubated at 37°C: T47D (Cat no: HTB-133), MDA-MB-231 (Cat no: HTB-26), Caki-1 (Cat no: HTB-46), ACHN (Cat no: CRL-1611), A498 (Cat no: HTB-44), and 786-O (Cat no: CRL-1932). T47D cells were grown in RPMI-1640 (ATCC, Cat no: 30-2001) with 0.2 U/mL insulin (Life Technologies, Cat no: 12585-014) and 10% FBS (ATCC, Cat no: 30-2020). MDA-MB-231 cells were grown in Leibovitz’s L-15 Medium (ATCC, Cat no: 30-2008) with 10% FBS. Caki-1 cells were grown in McCoy’s 5a Modified Medium (ATCC, Cat no: 30-2007) with 10% FBS. ACHN and A498 cells were grown in Eagle’s Minimum Essential Medium (ATCC, Cat no: 30-2003) with 10% FBS. 786-0 cells were grown in RPMI-1640 with 10% FBS.

### Quantification of SULT1A1 mRNA Expression by RT-qPCR

In triplicates, cells were plated in their designated complete media (described above) and treated with FIS103 at the concentrations and times indicated. Total RNA was isolated via Direct-zol RNA miniprep (Zymo Research, Cat no: R2050-1-200). 1ug of RNA was converted into cDNA using AffinityScript qPCR cDNA synthesis kit (AffinityScript, Cat no: ST600559). SULT1A1 gene specific primers and SYBR Green Mix were used for RT-qPCR on a StepOnePlus Real-Time PCR System (Applied Biosystems, Cat no: 4376600). SULT1A1 expression was normalized against 18S mRNA levels via Delta C_t_ method.

### Cell Viability Assays

6×10^3^ cells were plated in 96 well plates in designated complete media (described in Cell Lines and Media). After 1 day, cells were treated with indicated concentrations of FIS103 (50 nM, 100 nM, 250 nM, 500 nM, and 1 μM) or vehicle (DMSO; Sigma, Cat no: 67-68-5) for 72 hours; then CellTiter-Glo 2.0 Reagent was added to each well (CellTiter-Glo 2.0 Cell Viability Assay kit; Promega, Cat no: G9242) for a minimum of 10 min. Signal intensity was measured via luminescent plate reader. Compound-treated sample viability was normalized to the vehicle control.

### SULT1A1 Immunoblotting

Cell culture dishes were placed on ice, washed with ice-cold phosphate-buffered saline (PBS) (ThermoFisher, Cat no: 70011044), and lysed with ice-cold RIPA lysis buffer (Pierce, Cat no: 89900) supplemented with Halt™ protease inhibitors (ThermoFisher, Cat no: 87786). Cells were transferred into a pre-cooled microcentrifuge tube using a cold plastic cell scraper. Tubes were constantly agitated for 30 min at 4°C, centrifuged for 20 min at 12k rpm at 4°C, then placed on ice. The supernatant was transferred to a fresh tube on ice. An aliquot of cell lysate was taken to measure protein concentration via Rapid Gold BCA Protein Assay Kit (Pierce, Cat no: A53225). 4x Laemmli sample buffer (Bio-rad, Cat no: 1610747) was added to 20-30 μg cell lysate and boiled at 98°C for 5 min to denature the proteins. Equal amounts of protein were loaded into the wells of 4-20% TGX SDS-Page gels (Bio-rad, Cat no: 4561096); the gel was run for 1-2 hours at 120V in Tris Glycine SDS-Page Running Buffer (Bio-rad, Cat no: 1610732). Gel was transferred to iBlot Transfer Stack PVDF membrane (ThermoFisher, Cat no: IB401002) using the iBlotTM 2 Dry Blotting System (ThermoFisher, Cat no: IB21001). The membrane was cut at appropriate size markers and blocked for 1 hour at room temperature or overnight at 4°C using EveryBlot Blocking Buffer (Bio-rad, Cat no: 12010020). The membrane was incubated with SULT1A1 primary antibody (R&D Systems, Cat no: MAB5546) diluted in blocking buffer overnight at 4°C. The membrane was washed in PBS (3x, 5 min each) and then incubated with secondary antibody (goat anti-mouse IgG; BioRad, Cat no: 172-1011) diluted in blocking buffer for 1 hr at room temperature. Then, the same PBS washing process was repeated, and the immunoblots were developed using Odessey Fc Imaging System (LI-COR Biosciences). β-actin was measured as the loading control (Cell Signaling, Cat no: 8H10D10).

### Development of Stable Aqueous Formulation for FIS103

Formulation studies were conducted by the Southwest Research Institute (SwRI). Cyclodextrin (CD) complexes were purchased from Sigma-Aldrich and Fisher Scientific: 2-Hydroxypropyl-β-cyclodextrin (HPβCD), Sigma-Aldrich (Cat no: H107-5G); methyl-β-cyclodextrin (MβCD), Fisher Scientific (Cat no: AAJ6684706); Sulfobutylether-β-cyclodextrin (SBEβCD), Fisher Scientific (Cat no: NC1238671); and 2-Hydroxypropyl-γ-cyclodextrin (HPγCD), Sigma-Aldrich (Cat no: H125-5G-I). First, 450 mg of CD was dissolved into water, after which 5 mg of FIS103 was then dispersed. Regardless of concentration, samples for stability testing were prepared using the following steps at room temperature: 1) CD was weighed into a vial using a 5-place balance, 2) deionized water was added followed by 1-2 minutes of vortexing for MβCD solutions, or up to 8 minutes for HPβCD or HPγCD solutions, until the solutions were clear, 3) bubble formation during vortexing settled over 5-10 minutes (bath sonication was used, when necessary, to accelerate degassing), 4) after warming to room temperature in a separate vial, FIS103 was massed out using a 5-place balance (CD solution and a small stir bar were added to the vial), and 5) the solutions were vortexed for 1-2 minutes, followed by stirring for up to 2 hours until clear. Samples for evaluating solubility of FIS103 were prepared by adding 200 μL increments of CD solution to 5 mg FIS103. After each 200 μL addition, the suspension/solution was vortexed for 1-2 minutes and observed. If precipitate remained, an additional 200 μL was added, which was repeated until clarity was achieved. Stability was first evaluated at 2-8°C and room temperature (∼25°C). FIS103 was tested dry, in water, and in PBS. In the absence of CDs, 30% acetonitrile was used to solubilize FIS103 in aqueous systems for analysis using an Agilent 6140 liquid chromatography mass spectrometry (LC/MS) single quad system with a Kromasil C18 column (5 μm, 100Å, 30 × 4.60 mm). FIS103 levels at time 0 (T0) for each experiment was used to establish the 100% mark. Subsequent time points were compared to the T0 value to establish the percent recovered.

### *In Vivo* Efficacy Study of FIS103 in Subcutaneous A498 Tumor-Bearing Athymic Nude Mice

Studies were conducted by the CRO, Rincon Bio. A498 cells were previously purchased through ATCC (Cat no: HTB-44) and expanded for low passage aliquots in liquid nitrogen. Cells were thawed and cultured in RPMI + L-Glutamine, supplemented with 10% FBS, 1% Penicillin/Streptomycin solution, and 0.2% normocin (Invivogen), in a humidified incubator at 37°C and 5% CO_2_. Cells were split at 80% confluency and harvested for implantation at 70% confluency. Female homozygous NU/J mice (6-week old) were procured through Jackson Laboratory (Strain 002019). On implantation day, A498 cells were trypsinized and allowed to detach from flasks. Trypsin was then neutralized with complete media and cells were centrifuged at 400 x g. Media was aspirated and cells were washed with PBS without Ca^2+^ or Mg^2+^. Cells were resuspended in RPMI (non-supplemented) at 1×10^7^ cells/mL. 100 μL was injected into the right hind flank of each animal (a total of 1×10^6^ cells). Tumors were measured throughout the study by digital calipers in two dimensions, and the volume was calculated using the formula: Tumor Volume(mm^3^) = 0.5 x (*w*^2^ x *l*), where *w* = width and *l* = length, in mm, of the tumor.

### Analyzing FIS103 Treated SD Rats for Liver Toxicity

Sprague Dawley (SD) rats were treated with 25 mg/kg FIS103 or vehicle (DMSO) once daily via IP injection for 15 days. Serum samples and the rat livers were harvested at study conclusion to 1) analyze serum levels of liver enzymes alanine aminotransferase (ALT) and aspartate aminotransferase (AST) and 2) to conduct histopathology analyses of liver tissue sections. Liver enzyme levels were measured using ALT Colorimetric Activity Assay Kit (Cat no: 700260) and ASP Colorimetric Activity Assay Kit (Cat no: 701640), following manufacturer’s protocol. The liver tissue sections were analyzed by an experienced, board-certified pathologist in the Department of Anatomical Pathology at University of Texas MD Anderson Cancer Center.

## Results

### SULT1A1 as a Target for Cancer Therapeutics

SULT1A1 is a known inducer of carcinogenesis and a negative prognostic biomarker for many cancers, including RCC, low grade glioma, and uveal melanoma (Figure 1A)[38]. Thus, developing compounds that selectively target these aggressive cancer subtypes is beneficial. We examined two potential issues with using overexpression of SULT1A1 as a target for cancer therapeutics: (i) mutational variability within the SULT1A1 gene that may have functional implications; and (ii) homogeneity of SULT1A1 expression within the tumor. Consequently, we chose different RCC subtypes for analysis. We analyzed the TCGA PanCancer Atlas (Figure 1B) and the Genentech 2014 studies (Figure 1C). We discovered that SULT1A1 mutation can occur, but is very rare in RCC.

### Developing a Novel N-Benzyl Indole Carbinol (N-BIC) Analog

SULT1A1 bioactivates certain anticancer drugs like tamoxifen, therapy used to prevent or treat breast cancer. Several academic groups have assessed SULT1A1 as a TAE to activate other prodrugs. SULT1A1 is hypothesized to use its sulfotransferase activity to convert N-Benzyl Indole Carbinols (N-BICs), a promising class of chemical compounds, from their original inactive form (prodrug) to an electrophile active form. This active form has potent, non-specific alkylating agent properties leading to rapid cell death.

To develop more effective antitumor N-BIC analogs, we started a medicinal chemistry analytical campaign with scientists at American Biochemicals. We used *in silico* prediction software to generate new chemical analog compound classes (Figure 2B) based on the N-BIC core molecule (Figure 2A). Substituting an indole with an indazole ring as in compounds classes (1) and (3) was predicted to increase solubility. Similarly, substituting the terminal benzene ring with a pyridine (compounds (2) and (3)) was predicted to improve compound solubility.

We then did further *in silico* analysis and molecular designing with JRF International. We targeted a specific area of the parent molecule away from the indole for modifications. The computational analysis was based on *in silico* analyses of each analog’s solubility, predicted volume of distribution, and native pH level. Of the 20 specific chemical analogs generated with the best predicted performances across the metrices, FIS101, FIS102, and FIS103 were selected for synthesis, based on the metrics from the *in silico* analysis, i.e., best predicted solubility without compromising cell permeability (Figure 2C). The synthetic schemes had good yields, ranging from 60 to 80%. Analytical purity was measured by LCMS and storage stability was examined. Overall, FIS103, a substituted indole derivative, had better synthetic yield and higher stability than the other 2 compounds (Figure 2D).

Finally, FIS103 was tested for cell killing activity *in vitro*. RT-qPCR results show that on an mRNA level, the expression of SULT1A1 in T47D cells lines is 3-4 times greater than in MDA-MB-231 cell lines (Figure 3A). Cell viability assays revealed that FIS103 was most potent in killing T47D cells compared to the SULT1A1 low MDA-MB-231, reasserting SULT1A1-dependency (Figure 3B). It was further observed that FIS103 activity on T47D cells was concentration dose-dependent (Figure 3C). FIS103 was therefore selected for further experiments.

**Figure 3.**
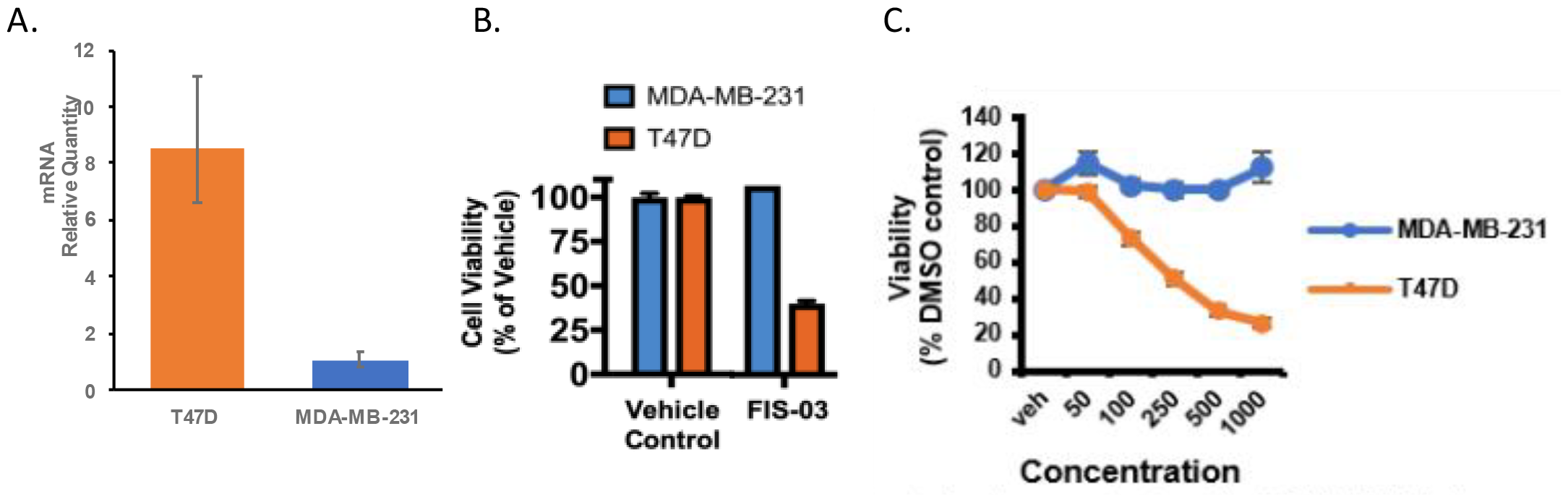
FIS103 induced cytotoxicity is dependent on SULT1A1 expression. A) SULT1A1 mRNA levels in SULT1A1 positive (T47D) and SULT1A1 negative (MDA-MB-231) cell lines. B) FIS03 demonstrated differential cell killing (cell viability assay) in T47D but not MDA-MB-231 cells, showing SULT1A1 dependency. C) The differential cytotoxic effect of FIS103 was dose dependent (concentration is nM). DMSO was used as vehicle control.

### Predicted Direct Interaction of FIS103 with SULT1A1

SULT1A1 is hypothesized to use its sulfotransferase activity to convert N-BIC compounds to active electrophiles that covalently alkylate proteins in cells, resulting in cell death (Figure 4A)[30]. We used *in silico* analysis to test whether this mechanism of action could explain the activity of FIS103. We collaborated with ACT to build a consensus high resolution crystal structure of SULT1A1 protein from publicly available PDB structures (IDs: 2D06 and 1LS6). When superimposed, the conformational differences at substrate binding sites among the different crystal structures fell under two clusters of conformations, of which PDB IDs 2D06 and 1LS6 were representative members. A bulk hydrophobic cavity of Phe142, Phe247, Phe24, Phe84, and Phe76 comprises SULT1A1’s substrate binding site; with Lys106 and His108 also participating in ligand interactions. Comparative analysis suggested that the Tyr240, Phe76 and Phe247 residues are displaced in PDB 2D06 because of the larger estradiol ligand. Both structures were selected for further analysis (Figure 4B).

**Figure 4.**
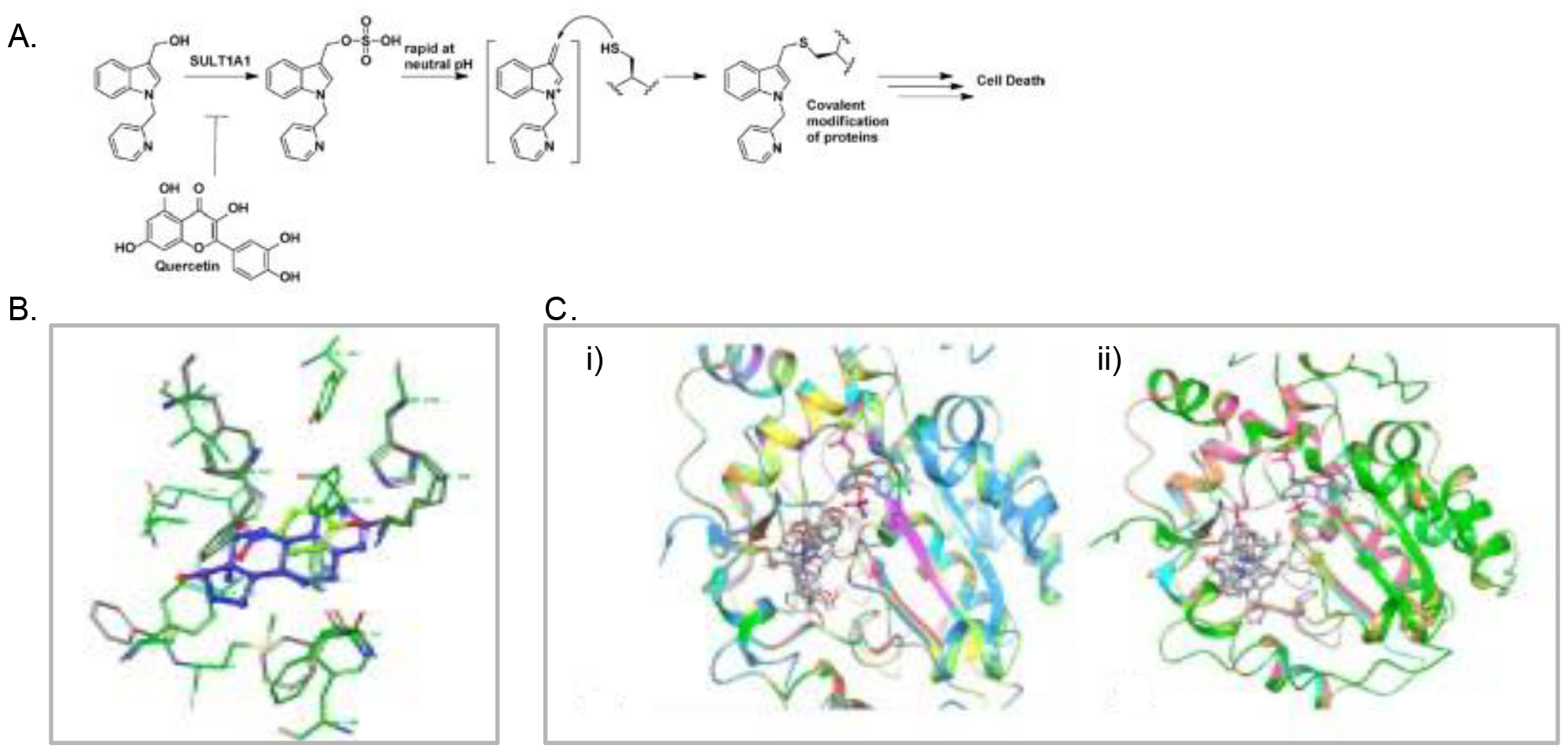
*In silico* analyses suggest FIS103 interacts with SULT1A1 in a similar fashion to N-BIC compounds. A) Proposed mechanism of action of N-BIC compounds after activation by SULT1A1 enzyme[30]. B) Aligned and superimposed representative structures of SULT1A1: 1LS6 (green) and 2D06 (grey). C) Cluster representative structure of FIS103 during 100ns MD simulation with i) 2D06 and ii) ILS6.

To generate a binding site model for FIS103 and compare the binding mode to that of p-nitrophenol or estradiol, FIS103 was docked at the defined grid using the automated SP mode of Glide. The docking outputs predicted that FIS103 binding at the SULT1A1 protein is very similar to p-nitrophenol or estradiol. To further confirm this, the top scoring docking models for 2D06 and 1LS6 was submitted to short 100 ns MD simulations using Gromacs v2020. The binding positions were well converged, as assessed by the time evolution of backbone atoms’ root mean square deviation (RMSD) from the starting structure. The RMSD analysis indicated that the ligand stays at the binding site during the simulation, but keeps reorienting. The lowest energy conformation of FIS103 aligns with co-crystallized conformation of estradiol and p-nitrophenol with 2D06 and 1LS6 (Figure 4C). Therefore, FIS103 likely uses the same mechanism of action with SULT1A1 as other N-BIC compounds.

### Improving the stability of FIS103

Despite a well-defined synthetic pathway, high yield, good solubility, and targeted cell killing, the stability of FIS103 in solution required optimization for further development as a lead compound. We evaluated aqueous stabilization strategies of FIS103 with SwRI, a leader in drug encapsulation and formulations. We tested three cyclodextrins (CDs) used for solubilization and stabilization in research and commercial products: methyl-β-cyclodextrin (MβCD), (2-Hydroxypropyl)-β-cyclodextrin (HPβCD), and (2-Hydroxypropyl)-γ-cyclodextrin (HPγCD) [42]. Concentration, pH, and temperature were varied to optimize formulation and stability. FIS103 solubility varies with different CDs, and is inversely correlated with CD concentrations (Supplemental Table S1).

Stability was evaluated at 2-8°C and room temperature. FIS103 was tested dry, solubilized in water, or in PBS (supplemented with 30% acetonitrile where CDs were absent) and CDs. Drug concentration at T0 was used as 100% to establish percent recovered (Supplemental Figure S1A). While stability of neat FIS103 is good at 2-8 °C and room temperature, the CDs improve stability further. The >100% recovery could be attributed to continued dissolution of FIS103 not captured at Time 0. The same solutions were heated to 40 °C and 60°C for 24 hours. Higher temperatures begin to show differences in performance between the CDs, with preference in the following order: MβCD > HPβCD > SBEβCD > HPγCD.

The first two were selected for further stability studies of FIS103 performed using 15 mM and 17.5 mM concentrations in 50 and 200 mg/mL PBS solutions of MβCD and HPβCD at 2-8 °C and room temperature for 7 weeks with MβCD (Supplementary Figure S1Bi) and 5 weeks with HPβCD (Supplementary Figure S1Bii). A 12-16% loss is observed after 7 weeks for MβCD for the two samples at room temperature; the two samples at 2-8 °C show an increase in recovery. For HPβCD, the decrease for the 17.5 mM solution at room temperature is slightly greater (decrease of 17% recovered drug). MβCD offers slightly better stability, but HPβCD has greater FIS103 dissolution capacity. Thus, FIS103 / HPβCD was chosen for further studies.

### *In Vitro* Efficacy Studies for FIS103 in Cancer Cells

Although FIS103’s dependency on SULT1A1 expression makes it efficacious in several tumor types, we chose a single cancer type, RCC, for consistency. Though 9% of RCC patients have SULT1A1 overexpression (Table 1), SULT1A1 expression in normal kidney tubules complicates targeting RCC with FIS103 *in vivo*. We evaluated FIS103 antitumor activity (50 nM to 1 μM) in 4 RCC cell lines whose SULT1A1 expression is high (A498 and Caki-1) or low (786-O and ACHN) (Figure 5A) based on protein and relative mRNA expression. Cell viability was measured after 72hr. A498, with the highest SULT1A1 expression, was the best responder to the FIS103 treatment - concentrations as low as 50 nM had efficient cell killing. Caki-1 was the other responder, whereas 786-O and ACHN were unaffected by the FIS103 treatment. The result clearly demonstrated FIS103 potency in SULT1A1 overexpressing cells and no activity in SULT1A1 low cells (Figure 5B). FIS103 was effective in cells derived from a primary tumor (A498) and from metastasized tumor (Caki-1). Cell viability assays confirmed FIS103 does not cause toxicity in concentrations as high as 1 μM for SULT1A1 low cells, 786-O and ACHN (Figure 5C). Conversely, the NSC-743380 molecule (parental compound to FIS103) caused cell death in low SULT1A1 expressing cells, MDA-MB-231, – indicating FIS103’s enhanced efficacy and reduced toxicity (Figure 5D).

**Figure 5.**
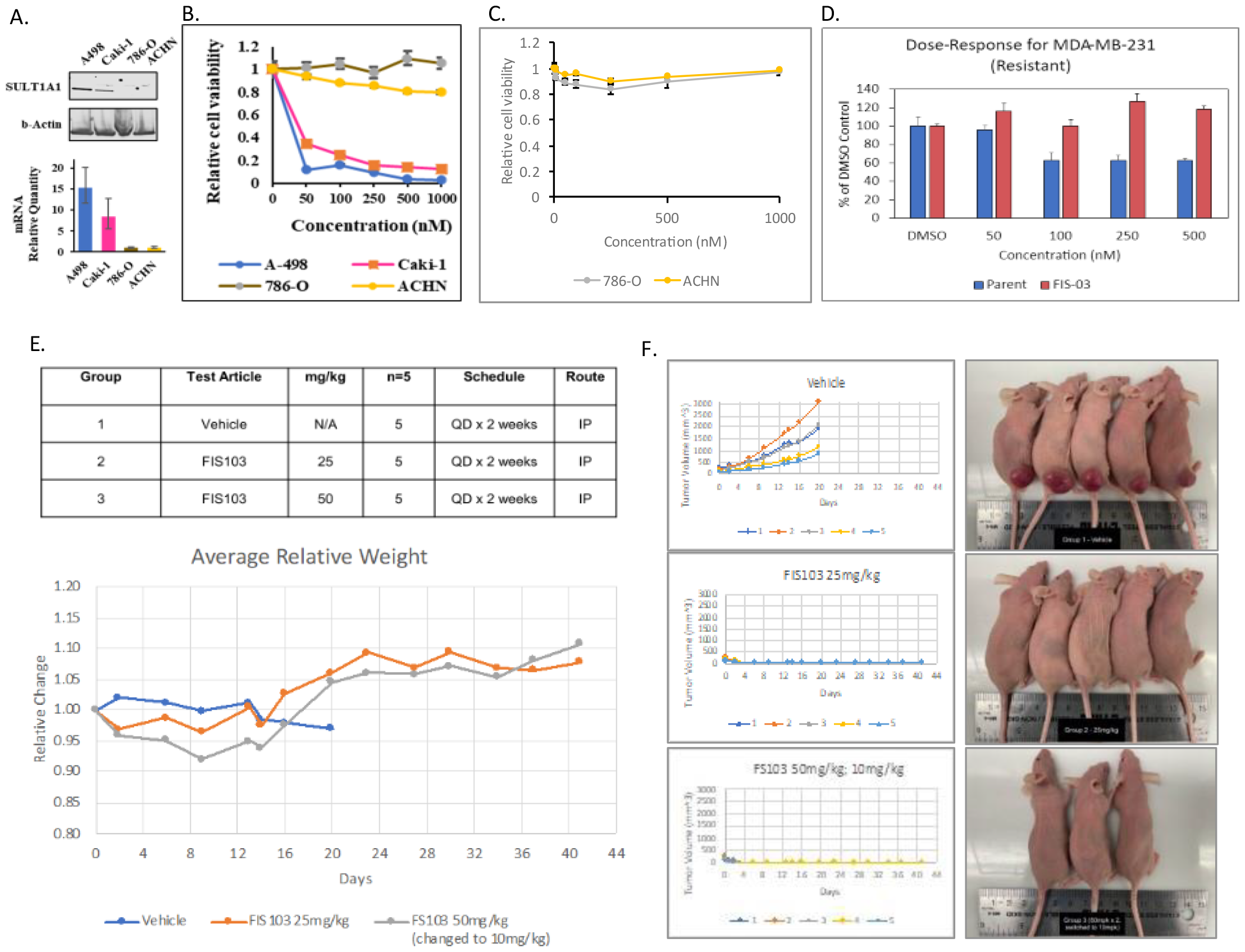
*In vitro* and *in vivo* efficacy studies for FIS103 in RCC models. A) SULT1A1 mRNA and protein expression in RCC SULT1A1 high- and SULT1A1 low-expressing cell lines. B) Cell viability assay demonstrating FIS103 treatment in SULT1A1 high-expression RCC cells. C) Cell viability assay demonstrating lack of toxicity of FIS103 up to 1 uM on cells with low SULT1A1 expression. D) Cell viability assay demonstrating lack of toxicity of FIS103 versus the parental compound (NSC-743380) in SULT1A1 non-expressing cells (MDA-MB-231). E) 6-week old NU/J mice were injected with 10^6^ A498 cells to allow tumor growth for 20 days before intraperitoneal injection of FIS103 at indicated concentrations for 14 days (once daily). Test groups for mice (table) and relative weights by treatment group (blue, vehicle; orange, 25 mg/kg; grey, 50 mg/kg then 10 mg/kg) over the course of the 41-day study. F) Tumor volume was measured over the course of the study (left). Control group tumors grew for 20 days and were then sacrificed due to excess tumor burden. Treatment groups were started at 24 mg/kg and 50 mg/kg doses. Mice getting 50 mg/kg showed signs of toxicity, and 2 died. The rest of this group were given a 3 day drug holiday and then treated at 10 mg/kg. All mice in the treatment groups had tumors that became non-detectable after 14 days and remained absent through study conclusion. Representative photos of the mice in each group are shown (right).

### *In Vivo* Studies for FIS103 in an RCC CDX Model

Evaluation of anti-tumor efficacy of FIS103 was done in an athymic RCC mouse model. Briefly, ∼10^6^ A498 cells/mouse were subcutaneously injected into the right hind flank of 6 weeks old female homozygous NU/J mice. Once the tumor size measured ∼93-278mm^3^, the mice were treated intraperitoneally with FIS103(vehicle, 25 mg/kg, or 50 mg/kg once daily for 14 days). Two days after treatment, the mice treated with 50 mg/kg were not tolerating FIS103 well, showing lethargy and 10% body weight loss; and 2/5 mice in that cohort succumbed from treatment. Treatment in this group was stopped for three days for drug washout, followed by dose reduction to 10 mg/kg. This group fully recovered - after acclimating, both 10 and 25 mg/kg treatment groups appeared to tolerate the drug and dosing schedule for the rest of the study.

The mice in the vehicle control group initially exhibited stable weight, but slowly lost weight (Figure 5E), likely from growing tumor burden. The 25 mg/kg group lost about 4% body weight in a few days but fully recovered. Tumors in the control group kept growing for 20 days; thus, the control group was sacrificed due to excess tumor burden. Remarkably, tumors in FIS103 treatment group remained small or regressed, becoming non-detectable after 14 days – tumors did not reappear through study conclusion (day 41) even after treatment concluded (Figure 5F).

### FIS103 Demonstrates Favorable Liver Toxicity Profile

SULT1A1 expression is higher in the liver and previous N-BIC compounds had shown off target toxicities, it was important to establish the liver toxicity profile of FIS103. Furthermore, most drugs that fail in preclinical/clinical development is due to toxic adverse events in the liver. Thus, healthy SD rats were treated with FIS103 or vehicle (DMSO) for 15 days (once/day, IP). The rat livers were harvested at study conclusion for tissue sectioning and histopathology analyses by an experienced pathologist. Additionally, in a repeat experiment, blood serum levels were harvested to analyze liver enzyme levels (alanine aminotransferase, ALT; aspartate aminotransferase, AST) as elevated serum levels for these enzymes can indicate liver toxicity. No pathological alterations were observed in the SD rat livers after FIS103 treatment (Figure 6A). Pathologist analysis indicated 1) liver architecture is intact, 2) no underlying fibrosis noted, 3) no signs of portal or lobular inflammation, which is commonly associated with toxic drugs and hepatocyte necrosis, and 4) no evidence of hemorrhage, necrosis, or steatosis, which is associated with chronic drug use liver toxicities. Serum levels of liver enzymes ALT and AST remained in the normal / healthy range in SD rats after 15 days of FIS103 treatment (Figure 6B).

**Figure 6:**
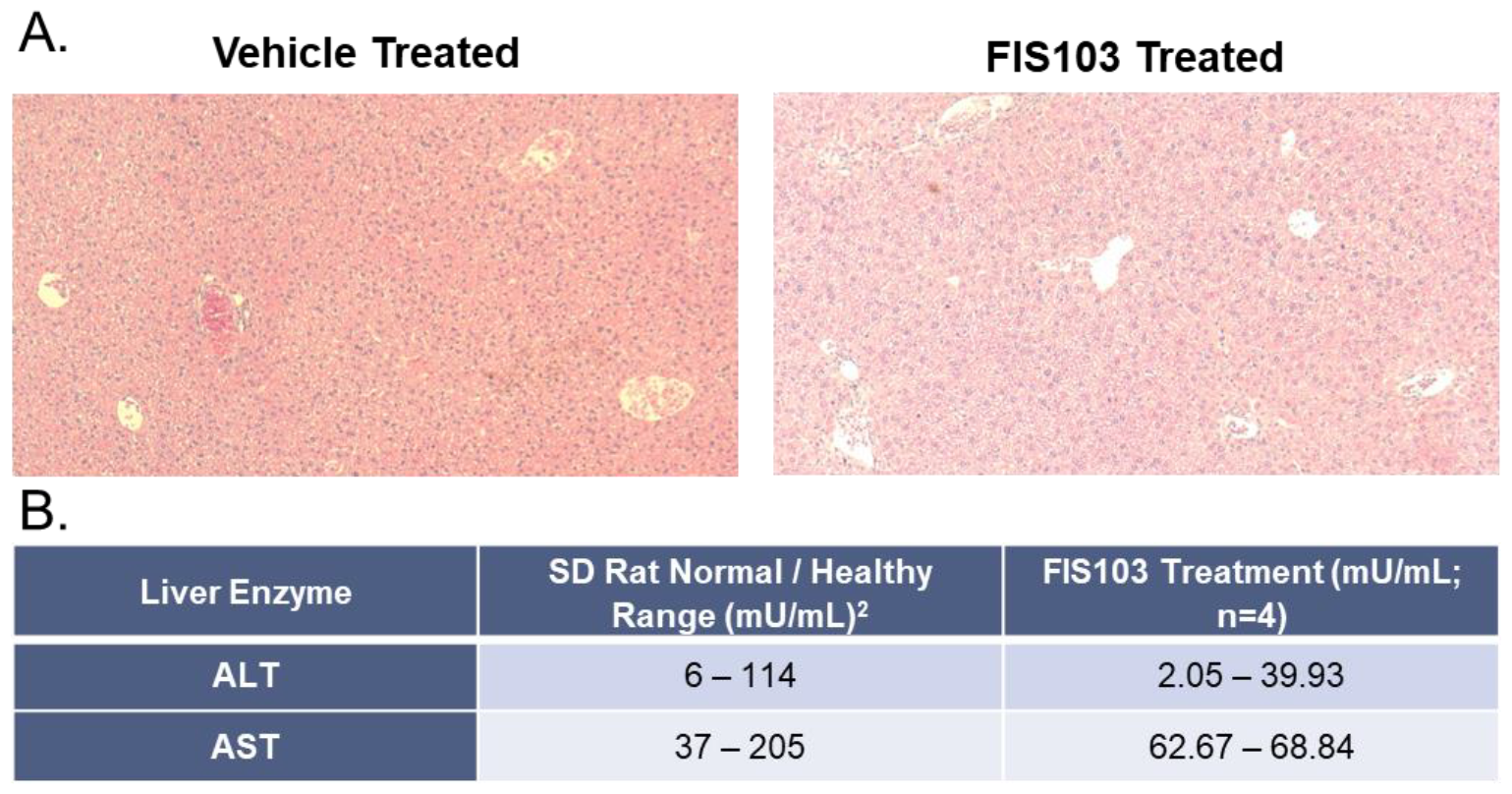
FIS103 displays a promising preliminary liver toxicity profile in SD rats. SD rats were treated with FIS103 (25 mg/kg, once daily, IP) or vehicle (DMSO, once daily, IP) for 15 days before harvesting the livers for tissue sectioning or harvesting serum to analyze liver enzyme levels (two repeat separate experiments). A) Representative images of liver tissue sections from FIS103 or vehicle treated SD rats. B) Serum levels of liver enzymes ALT and AST in FIS103 treated rats as compared to what is considered the normal healthy range for SD rats.

## Discussion

Targeted anticancer therapies capitalize on extracellular proteins found in cancer cells using antibody-based approaches (e.g., monoclonal antibodies or antibody drug conjugates) or engineered immune cells (e.g., CAR-T)[43-45]. Recently, the increasing application of proteomics and better understanding of subcellular processes in malignant cells has paved the way for chemotherapeutics with site-specific delivery of cytotoxic anticancer agents, coupling the advantages of both approaches[46-48]. Our lead compound FIS103 has a potentially superior mechanism of action versus existing chemotherapeutics considering its high specificity towards cancerous cells with SULT1A1 overexpression. Since SULT1A1 is lowly expressed systemically outside the gastrointestinal tract, parenteral administration may avoid most adverse effects. A simple companion diagnostic (CDx) screen for SULT1A1 (mRNA or protein) in a tumor biopsy or circulating tumor cells, would allow for identification and inclusion of patients more likely to respond.

Kidney and renal pelvis cancer is sixth most common for men, and eighth for women in the United States[49]. RCC is the most frequent renal cancer (∼85% of all renal malignancies)[50, 51]. The majority of RCC cases present at localized stage (∼65%) with relatively good prognosis. Unfortunately, one third of patients treated for localized disease will experience metastatic relapse at distal sites[52]. The 25%-30% of RCC cases who present with metastatic disease (mRCC) have poorer outcomes, particularly with distant metastasis (five-year survival rate of 12%)[53]. Thus, over half of patients diagnosed with RCC will require systemic therapy either as adjuvant therapy for localized patients with high-risk of recurrence or as first-line therapy for patients with mRCC[53, 54]. RCC does not respond well to conventional chemotherapy[55, 56]. Targeted therapy including small molecule tyrosine kinase inhibitors (sorafenib, sunitinib, and pazopanib), monoclonal antibody targeting VEGF (bevacizumab), and mTOR inhibitors (temsirolimus) have become first-line therapy for mRCC[51-53]. More recently, the checkpoint inhibitors (nivolumaband and pembrolizumab) have been approved for mRCC patients[57, 58]. Even though approved targeted therapies have improved survival for mRCC patients compared to the traditional approach, survival rate for mRCC patients is still poor, highlighting the urgent need for novel targeted approaches. Additionally, given the genetic diversity of RCC, there is a clear unmet need for targeted therapeutics for different subsets of RCC.

Our clinical candidate, FIS103, has demonstrated excellent *in vitro* and *in vivo* efficacy in SULT1A1-expressing RCC models and encouraging pharmaceutical properties including solubility and stability. FIS103 was effective against primary and metastatic RCC cells. Although SULT1A1 mutations can occur in RCC tumors, it is rare. SULT1A1 is homogenously expressed across RCC cancer cells, suggesting prodrugs activated by TAEs, such as FIS103, would be an effective targeted therapy. Future studies in patient-derived xenograft models of RCC, and other tumor types, are necessary to fully capture FIS103 preclinical *in vivo* efficacy.

SULT1A1 transfers a sulfate group at the hydroxyl position off the indole of N-BICs, which is rapidly cleaved at neutral pH within the cell, leaving a strong electrophile to interact with thiol groups (e.g., on cysteines). Therefore, this group non-specifically alkylates proteins with accessible thiol groups, resulting in shutdown of their activity, rapidly causing cell death. SULT1A1’s presence in the gut raises the possibility of severe dose limiting toxicity for N-BICs, although this may be largely avoided via parenteral administration. Preliminary toxicity studies by other groups using N-BICs have determined that, in mice, intravenous administration is tolerable at therapeutic doses[59]. Furthermore, mice in one study were dosed with NSC-743380 (parental compound) at 60 mg/kg[59]. Although we observed minor dose-dependent toxicity at 25 mg/kg, the lowest dose tested (10 mg/kg) resulted in an undetectable tumor at 14 days, suggesting a larger therapeutic window exists. Dose-ranging studies *in vivo* warrant further investigation to confirm FIS103 potential for similar antitumor effects. Nevertheless, potential for toxicity is a concern that needs to be carefully studied moving forward.

A second issue with N-BIC compounds is stability and solubility. N-BIC tool compounds are susceptible to inactivation by acid; thus, modifications are needed to improve stability[60]. Aqueous solubility needed for intravenous delivery is particularly critical for N-BICs because they are naturally lipophilic, and require substantial chemical modification / reformulation to achieve solubility.

Despite potential antitumor efficacy in different types of cancer, development of N-BIC analogs has lagged due to solubility / stability issues and SULT1A1-independent cytotoxicity, preventing their clinical advancement. We addressed these limitations through medicinal chemistry and formulation. FIS103 demonstrated enhanced stability, while maintaining SULT1A1-dependent potency. FIS103 is well tolerated in both *in vitro* and *in vivo* RCC models. Additionally, preliminary liver toxicity data in SD rats indicates that there is no visible morphologic differences between FIS103 or vehicle treated animals, along with liver enzyme serum levels falling within the healthy normal range for SD rats. This is promising data as the biggest limitation for N-BIC compounds is liver toxicity as SULT1A1 is higher expressed in this tissue. More extensive pharmacokinetics (PK) and toxicity studies of FIS103 are critical to determining treatment dose, safety pharmacology, and drug formulation.

Additionally, it is necessary to select an appropriate CDx. A CDx that can effectively classify patients based on expression of the predictive biomarker SULT1A1 is crucial to the success of FIS103. The most important criteria to select a CDx is accessibility and capacity to accurately predict patient response to treatment. Nucleic acid- (e.g., qRT-PCR, DNA microarray, or RNA-seq) and protein- (e.g., immunohistochemistry) based assays are both used in the clinic as CDx with targeted therapies[61]. To translate diagnostic variables into a clinical decision, it is critical to determine a cutoff point to stratify patients into distinct treatment groups[62]. The expression range of SULT1A1 that dictates sensitivity vs resistance to FIS103 remains to be determined.

Future work will evaluate PK and toxicity of FIS103, *in vivo* efficacy of FIS103 in multiple cancer models to demonstrate tissue-agnostic efficacy, and establish optimum dosage schedules. An appropriate CDx approach is needed to identify patient treatment groups. The development of this targeted chemotherapeutic has the potential to improve survival and quality of life of RCC patients. Furthermore, SULT1A1 has the potential to be a predictive biomarker and TAE in a variety of cancers. Therefore, FIS103 is a promising new targeted therapy with the potential to demonstrate tissue-agnostic efficacy in patients with SULT1A1-expressing tumors.

## Data Availability

All data produced in the present study are available upon reasonable request to the authors.

## Acknowledgements

Fannin would like to acknowledge 1) American Biochemicals and JRF International for contributions to the FIS103 MedChem Synthesis Campaign, 2) Texas Medical Center’s Accelerator for Cancer Therapeutics for i*n silico* prediction FIS103/SULT1A1 binding studies, and 3) Rincon Bio for *in vivo* FIS103 efficacy studies in RCC CDX models.

