## Supplemental Data for "FIS103, a Novel SULT1A1-dependent Prodrug, Demonstrates Potent Antitumor Activity in Renal Cell Carcinoma"

| FIS (mg) | CD | Amt (µL) | mM (w/ 450 mg/mL CD) | mM (w / 225 mg /mL CD) |
|---|---|---|---|---|
| 5 | HPγCD | 600 | 31.2* | 10.4 |
| 5 | HPβCD | 700 | 26.7 | 15.6 |
| 5 | SBME | 800 | 23.4 | 18.7 |
| 5 | MβCD | 1100 | 17.0 | 23.4 |

**Supplemental Table 1: FIS103 solubility in CD solutions.**

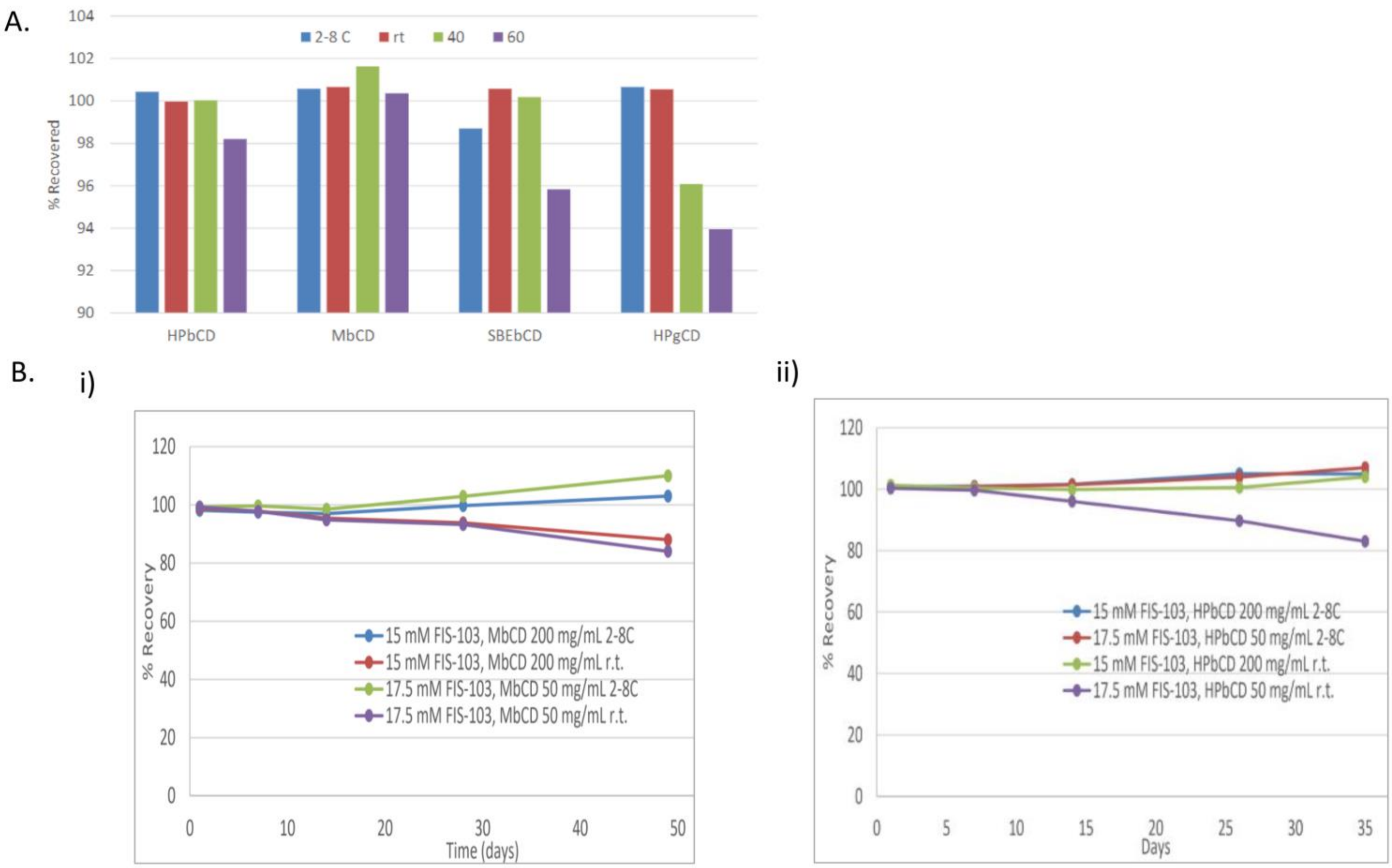


**Supplemental Figure S1. Formulation and stability assessment of FIS103.** A) Stability of 12.9 mM FIS103 in PBS at elevated temperatures for 24h in different cyclodextrins. The higher temperatures begin to show some differences in performance between the CDs, with preference in the following order: MβCD > HPβCD > SBEβCD > HPγCD. Concentration was measured using an Agilent 6140 LC/MS system with a Kromasil C18 column. B) Longer FIS103 stability studies in (i) MβCD or (ii) HPβCD over the course of 7 and 5 weeks, respectively. FIS103 concentrations were 15 mM and 17.5 mM in either 50 or 200 mg/mL CD-PBS solutions at 2-8oC or room temperature (r.t.).
